# Right ventricular pressure-strain relationship-derived myocardial work reflects contractility: validation with invasive pressure-volume analysis

**DOI:** 10.1101/2023.11.14.23298344

**Authors:** Bálint Károly Lakatos, Zvonimir Rako, Ádám Szijártó, Bruno R. Brito da Rocha, Manuel J. Richter, Alexandra Fábián, Henning Gall, Hossein A. Ghofrani, Nils Kremer, Werner Seeger, Daniel Zedler, Selin Yildiz, Athiththan Yogeswaran, Béla Merkely, Khodr Tello, Attila Kovács

**Author notes:** Corresponding author:* Attila Kovács MD, PhD, address: Varosmajor str. 68, Budapest, Hungary 1122. The first two (BKL and ZR) and last two authors (KT and AK) contributed equally to this manuscript.

## Abstract

**Background:** Three-dimensional (3D) echocardiography-derived right ventricular (RV) ejection fraction (EF) and global longitudinal strain (GLS) are valuable RV functional markers; nevertheless, they are substantially load-dependent. Global myocardial work index (GMWI) adjusts myocardial deformation to instantaneous pressure; therefore, it may better reflect contractility. Accordingly, we aimed to calculate RV GMWI using 3D echocardiography and examine its relationship with RV contractility’s gold standard invasive measurement.

**Methods:** Sixty patients (65±14 years, 65% female) with suspected or established pulmonary hypertension were enrolled. Detailed 3D echocardiography was performed to quantify RV EF and GLS using the ReVISION software. Patients underwent RV pressure-conductance catheterization to obtain the RV pressure curve and to assess contractility (end-systolic elastance - Ees) and ventriculo-arterial coupling (Ees/arterial elastance - Ees/Ea). RV GMWI was calculated using the RV GLS and the RV pressure trace curve.

**Results:** While neither RV EF (r=-0.143, p=0.275) nor GLS (r=-0.067, p=0.611) correlated with Ees, GMWI showed a strong correlation with it (r=0.669, p<0.001). In contrast, RV EF and GLS showed a relationship with Ees/Ea (RVEF: r=0.552; GLS: r=0.460, both p<0.001). By dividing the population based on the Reveal Lite 2 risk classification, EF and GLS showed a significant decrease only in the high-risk group (low vs. intermediate vs. high risk; EF: 48.8±6.6 vs. 44.0±9.4 vs. 35.0±7.8%, ANOVA p<0.001; GLS: 18.6±3.6 vs. 17.0±4.5 vs. 13.3±3.9%, ANOVA p<0.001), whereas GMWI already showed an increase in the intermediate group (550±267 vs. 831±361 vs. 797±265 mmHg%, ANOVA p<0.01).

**Conclusions:** RV EF and GLS reflect ventriculo-arterial coupling, while GMWI strongly correlates with contractility. RV GMWI may emerge as a useful clinical tool for risk stratification and follow-up in patients with pulmonary hypertension.

---

Myocardial contractility represents the intrinsic ability of the myocardium to shorten independently of loading conditions and, as such, is the target feature of ventricular performance for patient evaluations. In clinical practice, the assessment of myocardial function is most commonly performed using echocardiography with a constantly augmenting technical toolkit. Importantly, even advanced echocardiographic metrics, such as three-dimensional (3D) echocardiography-derived ejection fraction (EF) or global longitudinal strain (GLS), are heavily dependent on loading conditions; therefore, they can not be considered reliable markers of contractility (1). Increased afterload can significantly diminish the value of GLS despite maintained or even increased contractile function; thus, GLS rather reflects ventriculo-arterial coupling (1). To mitigate the afterload dependency of left ventricular (LV) GLS, the concept of myocardial work by adjusting GLS to the instantaneous LV pressure has been validated and subsequently introduced in clinical practice (2). Previous experimental work demonstrated that the global myocardial work index (GMWI) correlates with gold-standard pressure-volume analysis-derived measures of contractility in different hemodynamic overload conditions; however, EF or GLS is not (3). Importantly, the issue of load-dependency may culminate in the right side of the heart: right ventricular (RV) systolic performance is even more intensely exposed to alterations in afterload, yet the concept of myocardial work has not been thoroughly tested in its context. Available data are based on a two-dimensional (2D) echocardiography, which carries significant limitations due to the RV’s complex shape, contraction pattern, and hemodynamics, impeding an identical approach to the LV.

Accordingly, our present study aimed to calculate RV myocardial work using 3D echocardiography-derived RV GLS and examine its relationship with the gold-standard invasive measurement of RV contractility.

This research constitutes a post-hoc analysis of data collected from the EXERTION study (ClinicalTrials.gov Identifier: NCT04663217). The study adhered to the principles outlined in the Declaration of Helsinki and was approved by the local Ethics Committee of the Faculty of Medicine at the University of Giessen (Approval Number: 117/16). Written informed consent was obtained from all participating patients. Subjects were either undergoing an initial invasive diagnostic evaluation for suspected pulmonary hypertension or were already diagnosed with pulmonary hypertension. All patients underwent 3D echocardiography and RV pressure-conductance catheterization. Using the Reveal Lite 2 risk stratification score, low- (n=23), intermediate- (n=20), or high-risk (n=17) groups were identified. The technical details of the echocardiographic examinations and the RV pressure-conductance catheterization were described in detail previously (4). Briefly, 3D echocardiography datasets were analyzed using the 4D RV-Function 2 software (TomTec Imaging GmbH, Unterschleissheim, Germany) to calculate RV volumes and EF. To assess the 3D GLS, reconstructed 3D mesh models were analyzed with the ReVISION software (Argus Cognitive, Inc., Lebanon, NH, USA), employing a previously reported methodology (5). Subjects also underwent RV pressure-conductance catheterization (Inca, CD Leycom, Zoetermeer, The Netherlands); therefore, we measured RV systolic pressure (RVSP), and during preload manipulation by Valsalva-maneuver, arterial elastance (Ea) and end-systolic elastance (Ees - gold standard measure of contractility) were calculated. Using these measures, we also quantified ventriculo-arterial coupling (Ees/Ea).

In myocardial work analysis, we followed previously published principles (2). GLS curves and invasively-acquired RV pressure recordings were exported and analyzed. The isovolumetric phases were identified using the second derivative squared method by expert consensus reading on the RV pressure tracing. By inspecting the RV volume curve, isovolumetric phases were also identified by expert consensus reading, and corresponding time points were applied to the GLS curve. Due to the different temporal resolutions of the datasets, the timestamps of the pressure and strain tracings were normalized in each section, and strain values were interpolated for the timestamps of the RV pressure recording. Then, the four sections of each recording were concatenated, and pressure– strain loops were constructed and plotted. The instantaneous power was calculated by multiplying the strain rate (obtained by differentiating the strain curve) and the instantaneous RV pressure. GMWI was computed by integrating the power over time from the beginning of isovolumetric contraction until the end of isovolumetric relaxation.

The study included sixty patients, with an average age of 65±14 years, and 65% of whom were female.

RV EF (r=-0.143, p=0.275) and GLS (r=-0.067, p=0.611) did not correlate with Ees, but rather with Ees/Ea (RVEF: r=0.552, p<0.001; GLS: r=0.460, p<0.001). In contrast, GMWI inversely correlated with Ees/Ea (r=-0.439, p<0.001) but, importantly, showed a strong direct correlation with Ees (r=0.669, p<0.001).

By dividing the patients based on their Reveal Lite 2 risk classification, every 3D echocardiographic and invasive hemodynamic marker showed significant differences (Table 1). By post-hoc analysis, the low-risk group had comparable EF and GLS to intermediate patients, while GMWI was significantly higher in the latter. Ees/Ea significantly decreased in the intermediate group, while RVSP was higher. High-risk patients significantly differed in every measure compared to the low-risk group. By comparing intermediate and high risk, EF and GLS were significantly lower in the latter, while GMWI and the invasive hemodynamic measures did not differ (Table 1). Representative cases of each group are depicted in Figure 1.

**Table 1:**
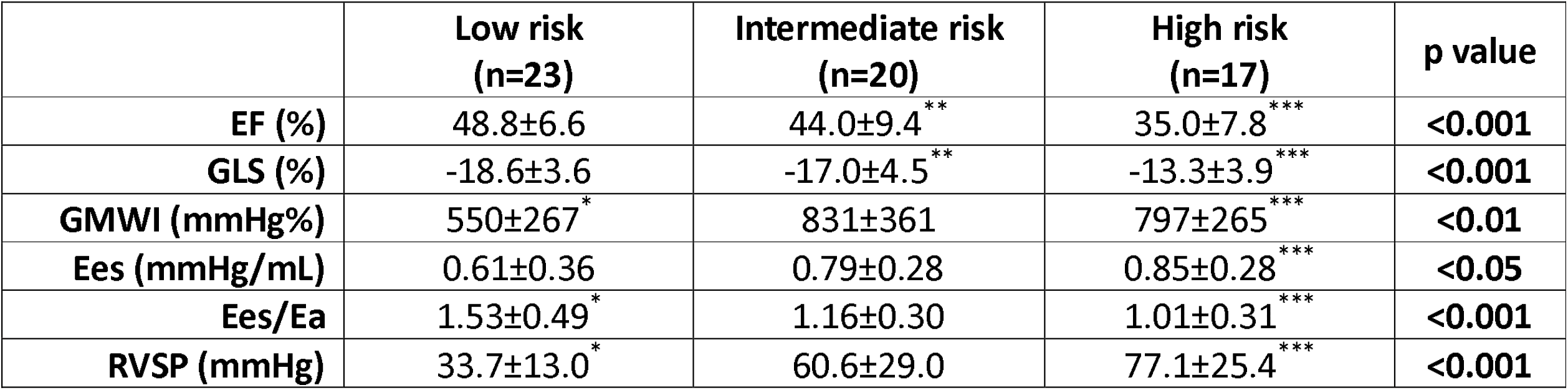
3D echocardiographic and invasive hemodynamic data of the patients by Reveal Lite 2 risk categories. Abbreviations: EF: ejection fraction; GLS: global longitudinal strain; GMWI: global myocardial work index; Ees: end-systolic elastance; Ees/Ea: ventriculo-arterial coupling; RVSP: right ventricular systolic pressure ^*^: low vs. intermediate p<0.05; ^**^: intermediate vs. high p<0.05; ^***^: low vs. high p<0.05

**Figure 1:**
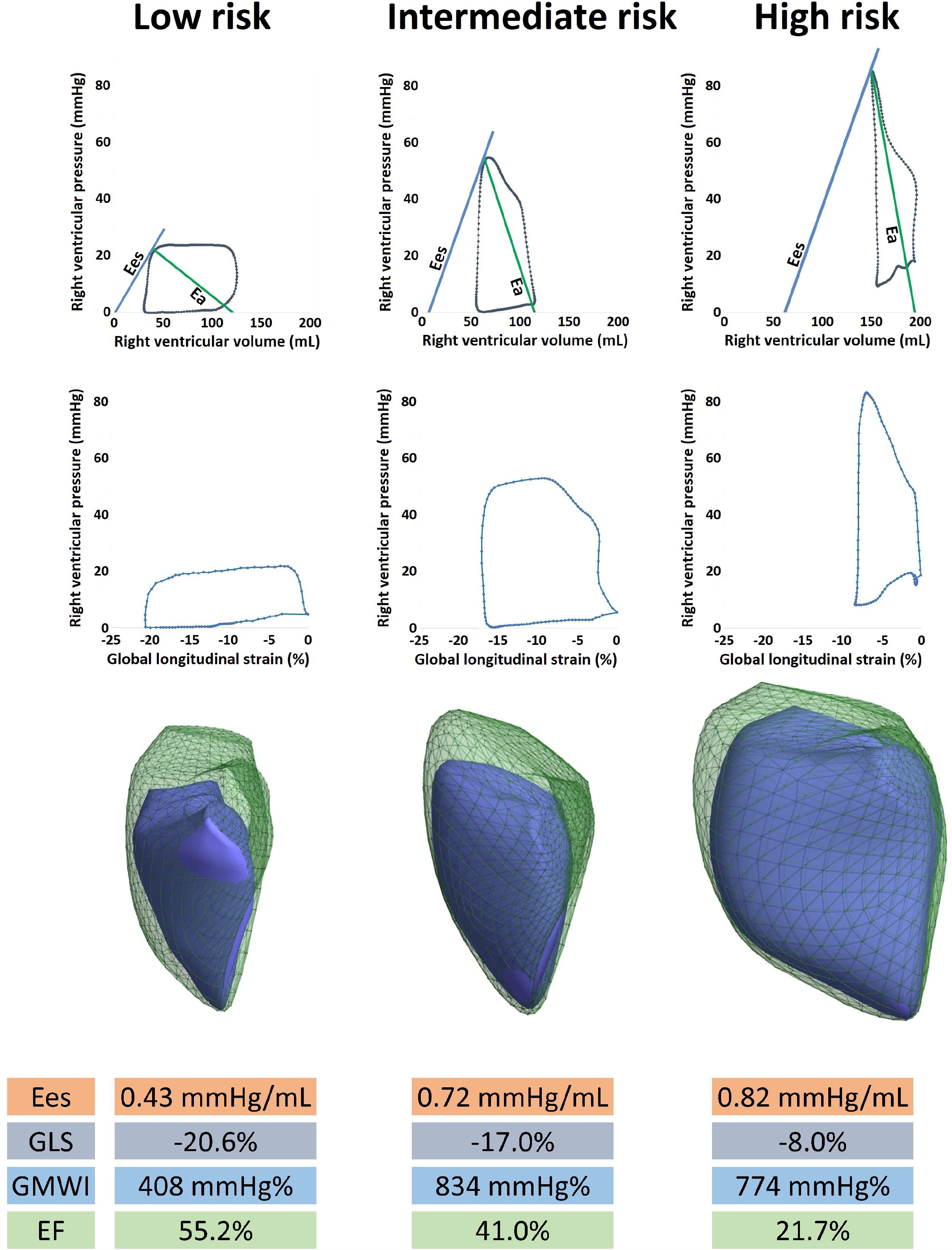
representative cases of each Reveal Lite 2 risk category. The low-risk patient demonstrates the lowest end-systolic elastance (Ees) and global myocardial work index while having maintained global longitudinal strain (GLS) and ejection fraction (EF). In the intermediate-risk patient, higher Ees and GMWI can be observed, while both GLS and EF show moderate impairment. In the high-risk patient, while Ees and GMWI are numerically comparable to the intermediate patient, EF and GLS are heavily deteriorated. Significant chamber dilation can also be observed as a sign of ventriculoarterial uncoupling (Ees/Ea=0.67).

Our findings indicate that similarly to the observations in the LV, RV EF and GLS reflect ventriculo-arterial coupling rather than myocardial contractility. On the other hand, the fusion of 3D echocardiography-derived GLS with instantaneous RV pressures allows the quantification of RV GMWI, which strongly correlates with the gold standard measure of RV contractility. GMWI is able to display the increased contractile state of the RV during the course of ventriculoarterial uncoupling, making it an appealing tool for more precise risk stratification and follow-up for patients with pulmonary hypertension. Further developments should aim at the accurate and non-invasive estimation of the individual RV pressure tracing to allow the everyday clinical use of the RV myocardial work concept.

## Data Availability

All data produced in the present study are available upon reasonable request to the authors

## Notes

*Disclosures and funding:* This study was funded by Deutsche Forschungsgesellschaft, Grant Agreement Number CRC 1213, Project B08. Project number RRF-2.3.1-21-2022-00003 has been implemented with support from the European Union. Dr. Kovács was supported by the Janos Bolyai Research Scholarship of the Hungarian Academy of Sciences. Dr Rako, Ms Yildiz, Mr da Rocha and Dr Kremer report nonfinancial support from the University of Giessen during the conduct of the study. Dr Yogeswaran reports nonfinancial support from the University of Giessen during the conduct of the study and personal fees from MSD outside the current study. Dr Lakatos, Mr. Szijártó, Dr Fábián, and Dr Kovács report personal fees from Argus Cognitive, Inc. during the conduct of the study. Dr Ghofrani reports nonfinancial support from the University of Giessen and grants from the German Research Foundation during the conduct of the study and personal fees from Bayer, Actelion, Pfizer, Merck, GSK and Takeda, grants and personal fees from Novartis, Bayer HealthCare and Encysive/Pfizer, and grants from Aires, the German Research Foundation, Excellence Cluster Cardiopulmonary Research, and the German Ministry of Education and Research outside the current study. Dr Seeger reports grants from the German Research Foundation and nonfinancial support from the University of Giessen during the conduct of the current study and personal fees from Pfizer and Bayer Pharma AG outside the submitted work. Dr Gall reports grants from the German Research Foundation and nonfinancial support from the University of Giessen during the conduct of the current study and personal fees from Actelion, AstraZeneca, Bayer, BMS, GSK, Janssen-Cilag, Lilly, MSD, Novartis, OMT, Pfizer, and United Therapeutics outside the current study. Dr Richter reports nonfinancial support from the University of Giessen during the conduct of the current study and research support from United Therapeutics and Bayer, speaker honoraria from Bayer, Actelion, Mundipharma, Roche, and OMT, and consultancy fees from Bayer outside the submitted work.

### Competing Interest Statement

Dr Rako, Ms Yildiz, Mr da Rocha and Dr Kremer report nonfinancial support from the University of Giessen during the conduct of the study. Dr Yogeswaran reports nonfinancial support from the University of Giessen during the conduct of the study and personal fees from MSD outside the current study. Dr Lakatos, Mr. Szijarto, Dr Fabian, and Dr Kovacs report personal fees from Argus Cognitive, Inc. during the conduct of the study. Dr Ghofrani reports nonfinancial support from the University of Giessen and grants from the German Research Foundation during the conduct of the study and personal fees from Bayer, Actelion, Pfizer, Merck, GSK and Takeda, grants and personal fees from Novartis, Bayer HealthCare and Encysive/Pfizer, and grants from Aires, the German Research Foundation, Excellence Cluster Cardiopulmonary Research, and the German Ministry of Education and Research outside the current study. Dr Seeger reports grants from the German Research Foundation and nonfinancial support from the University of Giessen during the conduct of the current study and personal fees from Pfizer and Bayer Pharma AG outside the submitted work. Dr Gall reports grants from the German Research Foundation and nonfinancial support from the University of Giessen during the conduct of the current study and personal fees from Actelion, AstraZeneca, Bayer, BMS, GSK, Janssen-Cilag, Lilly, MSD, Novartis, OMT, Pfizer, and United Therapeutics outside the current study. Dr Richter reports nonfinancial support from the University of Giessen during the conduct of the current study and research support from United Therapeutics and Bayer, speaker honoraria from Bayer, Actelion, Mundipharma, Roche, and OMT, and consultancy fees from Bayer outside the submitted work.

### Funding Statement

This study was funded by Deutsche Forschungsgesellschaft, Grant Agreement Number CRC 1213, Project B08. Project number RRF-2.3.1-21-2022-00003 has been implemented with support from the European Union. Dr. Kovacs was supported by the Janos Bolyai Research Scholarship of the Hungarian Academy of Sciences.

### Author Declarations

This research constitutes a post-hoc analysis of data collected from the EXERTION study (ClinicalTrials.gov Identifier: NCT04663217). The study adhered to the principles outlined in the Declaration of Helsinki and was approved by the local Ethics Committee of the Faculty of Medicine at the University of Giessen (Approval Number: 117/16).

